# Personalized Management of Septic Shock Guided by Multimodal Circulatory and Perfusion Monitoring: The PRISM Trial

**DOI:** 10.1101/2025.10.02.25337153

**Authors:** Athanasios Chalkias

**Affiliations:** Institute for Translational Medicine and Therapeutics, University of Pennsylvania Perelman School of Medicine, Philadelphia, PA, USA; Outcomes Research Consortium, Houston, TX, USA; Department of Critical Care Medicine, Tzaneio General Hospital, Piraeus, Greece

**Keywords:** septic shock, multimodal monitoring, hemodynamics, tissue perfusion, microcirculation, SOFA, SAPS II

## Abstract

**Background:** Sepsis-related organ dysfunction results from complex interactions between systemic hemodynamics, microcirculatory alterations, and cellular metabolic failure. Conventional resuscitation strategies guided by global parameters may miss persistent tissue hypoperfusion, a phenomenon termed “hemodynamic incoherence.” The PRISM trial was designed to determine whether individualized management guided by advanced multimodal circulatory and perfusion monitoring improves outcomes in septic shock.

**Methods:** The PRISM trial is a multicenter, randomized, controlled, open-label study with blinded outcome assessment. Adults with septic shock (Sepsis-3 criteria) are randomized (1:1) to structured multimodal monitoring versus standard care. The intervention integrates advanced systemic hemodynamic indices —including mean circulatory filling pressure analogue and other determinants of venous return, heart efficiency, cardiac power output, power efficiency, and volume efficiency— with a comprehensive perfusion panel (capillary refill time, mottling score, temperature gradients, lactate kinetics, central venous oxygen saturation, venous–arterial carbon dioxide difference, near-infrared spectroscopy-derived skeletal muscle tissue oxygen saturation, and arterial–interstitial glucose gradients). A predefined treatment algorithm links abnormal thresholds to therapeutic interventions. The primary endpoint is change in SOFA and SAPS II scores from baseline to 72 hours. Secondary endpoints include 28-day mortality, ICU and hospital length of stay, ventilator- and vasopressor-free days, lactate clearance, and safety outcomes.

**Discussion:** By combining advanced hemodynamic physiology with structured multimodal perfusion monitoring, the PRISM trial tests whether individualized, pathophysiology-guided resuscitation can overcome hemodynamic incoherence and improve patient-centered outcomes in septic shock.

## I. BACKGROUND

Sepsis-related organ dysfunction is caused by a dysregulated host response to infection and involves a variety of interrelated mechanisms, such as cellular injury, metabolic disruptions, coagulation abnormalities, and inflammatory reactions.^1,2^ In recent years, the focus of research on this pathophysiology has shifted from simply identifying the underlying etiology of the disease to examining physiological impairments at all biological levels, including changes in the microcirculation.^3^ The latter are crucial in the development of shock and organ failure and, in the majority of patients —if not all— occur before macrocirculatory alterations during the early phases of sepsis.^4-7^

Peripheral perfusion monitoring provides a severity measure that can evaluate hemodynamic incoherence and direct therapy, despite the fact that there are currently no reliable techniques for tracking the patient’s status.^8,9^ This method provides a visual assessment of tissue perfusion by monitoring blood flow in the peripheral microcirculation using physical examination, imaging procedures, and other techniques. Because peripheral perfusion monitoring is easy to access and non-invasive, its use has increased in recent years.

Although individual tests such as capillary refill time (CRT) have been evaluated in clinical practice,^10,11^ a comprehensive multimodal assessment of circulatory and perfusion dynamics has not yet been systematically implemented. The PRISM trial is designed to address this gap by integrating complementary monitoring approaches, including novel techniques such as skeletal muscle microvascular oxygenation (StO_2_). This multimodal strategy aims to provide a precise, real-time, and multidimensional evaluation of hemodynamic status, peripheral vascular reactivity, and tissue perfusion,^12–16^ thereby guiding personalized management of septic shock and enabling timely therapeutic interventions.

The PRISM trial is a randomized controlled trial (RCT) designed to determine whether a personalized management strategy guided by multimodal circulatory and perfusion monitoring improves outcomes in patients with septic shock.

## II. METHODS

### 1. Study design

This will be a multicenter, parallel group, randomized, controlled, open label trial with blinded outcome assessment. Critically ill patients admitted to the intensive care unit (ICU) will be eligible for inclusion. The study protocol will be approved by the Ethics Committee of Tzaneio General Hospital, Piraeus, Greece. The trial will be conducted in accordance with the principles of the Declaration of Helsinki and will be prospectively registered at ClinicalTrials.gov. Written informed consent will be obtained from each patient or, when necessary, from their legally authorized representative.

### 2. Objective

The primary objective is to determine whether multimodal circulatory- and perfusion-guided management reduces organ dysfunction, assessed as the change in SOFA and SAPS II scores from baseline to 72 hours, versus standard care.

### 3. Population

#### 3.1 Inclusion criteria

1. Age ≥ 18 years.
2. Admission to ICU with septic shock, defined by Sepsis-3 criteria: suspected or confirmed infection and vasopressor requirement to maintain mean arterial pressure (MAP) ≥ 65 mmHg despite adequate fluid resuscitation, and serum lactate > 2 mmol L^-1^.
3. Onset of septic shock ≤ 6 hours prior to randomization.
4. Expected to remain in the ICU ≥ 72 hours.
5. Informed consent obtained from the patient or legally authorized representative.

#### 3.2 Exclusion criteria

1. Age < 18 years.
2. Advance directives limiting escalation of care (comfort measures only).
3. Expected death within 24 hours regardless of treatment.
4. End-stage comorbidity likely to determine 28-day outcome (e.g., end-stage malignancy, Child-Pugh C) —to reduce outcome confounding from irreversible disease.
5. Local conditions interfering with peripheral perfusion monitoring or tissue oxygen saturation (StO_2_) sensors (e.g., severe peripheral vascular disease, limb amputation, extensive skin breakdown, burns, massive edema.
6. Contraindication to study monitoring devices (e.g., adhesive allergy, implanted device interference).
7. Active major hemorrhage or uncontrolled bleeding.
8. Primary cardiogenic shock or acute coronary syndrome as the dominant cause of shock.
9. Concurrent enrollment in another interventional trial with conflicting endpoints or interventions.
10. Prior enrollment in PRISM during the same hospitalization.
11. Vulnerable populations where consent cannot be obtained or protected per local regulations (e.g., prisoners), unless special provisions are made and approved by ethics boards.

### 4. Interventions

#### 4.1 Intervention arm

Patients in the intervention arm will undergo structured multimodal circulatory and peripheral perfusion assessments every 4 hours for 72 hours. A predefined treatment algorithm (PTA) will be followed. Algorithm steps (in order):

1. Assess fluid responsiveness using passive leg raise and/or dynamic indices. If fluid responsive → give 500 mL balanced crystalloid bolus over 20 min; reassess perfusion parameters and stop if improved.
2. If not fluid responsive or persistent abnormality despite fluid optimization → optimize MAP with vasopressor to individualized target (based on pre-admission MAP if known; otherwise default 65 mmHg), titrate slowly and reassess.
3. If cardiac dysfunction suspected (e.g., low CO or low LVOT-VTI) → consider inotrope (e.g., dobutamine) per local practice.
4. If sepsis-related, non-compensatory tachyarrhythmia → optimize with intravenous landiolol (see section 8: Data collection, monitoring, management, and schedule of assessments).
5. Evaluate potential sources of infection and expedite diagnostics/source control.
6. If StO_2_ very low and refractory → consider transfusion if hemoglobin below local transfusion threshold and clinician deems appropriate.
7. Reassess every four hours.

All therapeutic decisions remain under treating clinician control —the protocol is guidance, not mandatory orders. Deviations must be recorded. In addition to the interventions specified in the interventional arm, all patients will receive standard management in accordance with the Surviving Sepsis Campaign guidelines and institutional ICU protocols.^17,18^

#### 4.2 Control arm — Standard care

Patients will receive usual management according to the Surviving Sepsis Campaign guidelines and local ICU protocols.^17,18^ Treating clinicians will have access to conventional hemodynamic variables (e.g., MAP, heart rate, CVP, lactate) but will not be provided with the structured multimodal perfusion summaries or algorithm-based prompts from the intervention arm. Routine bedside assessments of perfusion (e.g., CRT) may be performed at the clinician’s discretion, but without protocolized treatment triggers.

### 5. Randomization and blinding

Randomization will be performed centrally through a secure, web-based system using permuted blocks, stratified by study site and baseline APACHE II category (≤15 vs. >15; allocation ratio 1:1). Given the nature of the intervention, blinding of treating clinicians is not feasible; therefore, the trial will be conducted in an open-label fashion. However, to minimize bias, outcome assessors —including those adjudicating SOFA and SAPS II scores and the trial statisticians— will remain blinded to treatment allocation.

### 6. Assessment of circulatory and perfusion dynamics

#### 6.1 Hemodynamic assessment

A patient-centered strategy will be applied to provide a precise, quantitative evaluation of each individual’s circulatory dynamics.

##### 6.1.1 General assessment

The internal jugular or subclavian vein will be cannulated with a triple-lumen central venous catheter that will be connected to another pressure transducer to measure central venous pressure (CVP) and central venous oxygen saturation (ScvO_2_). The radial artery will be cannulated and connected to a closed-circuit set and transduction system communicating with a monitor, allowing the direct measurement of systolic arterial pressure (SAP), diastolic arterial pressure (DAP) and MAP pressure.

Critical care echocardiography will be performed twice daily by a highly experienced echocardiographer who will be blinded to the patient’s identity and study sequence. Left ventricular ejection fraction (LEVF), left ventricular outflow tract (LVOT), and LVOT velocity time integral (LVOT-VTI) will be recorded among others. All echocardiographic parameters will be calculated from five measurements (regardless of the respiratory cycle) and analyzed retrospectively.

Before making each measurement, we will confirm that transducers are correctly leveled and zeroed,^19,20^ while the system’s dynamic response will be confirmed with fast-flush tests.^21^ Damping will be assessed via repeatedly performed visual inspections of the pressure waveform at the end of a fast flush test. Artifacts will be detected and removed when documented, and when measurements are out-of-range or SAP and DAP are similar or abruptly changed (≥40 mmHg decrease or increase within two minutes before and after measurement). The amount and type of fluids and the dose of vasopressors will be titrated to maintain a MAP level based on the patient preadmission levels via clinical judgment and dynamic/static tests.

##### 6.1.2 Determinants of venous return

The methods of the mean circulatory filling pressure (Pmcf) analogue (Pmca) and related values algorithm have been described in detail before.^5,22-24^ Briefly, based on a Guytonian model of the systemic circulation [CO = *V*R = (Pmcf − CVP) /RVR], an analogue of Pmcf can be derived using the mathematical model Pmca = (a × CVP) + (b × MAP) + (c × CO). In this formula, a and b are dimensionless constants (a + b = 1). Assuming a veno-arterial compliance ratio of 24:1, ‘a’ = 0.96 and ‘b’ = 0.04, reflecting the contribution of venous and arterial compartments, and ‘c’ is a combination of veno-arterial compliance ratio (=0.96) and venous compartment resistance (=SVR × 0.038), with the dimensions of resistance, and is based on a formula including age, height, and weight^25^:

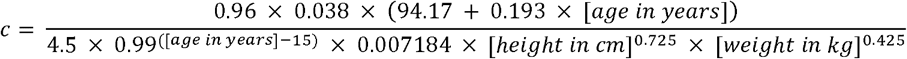

Driving pressure for venous return (VRdP) will be defined as the pressure difference between Pmca and CVP (VRdP = Pmca – CVP). Resistance to venous return (RVR) will be defined as the resistance downstream of Pmca to reflect resistance for venous return and will be calculated as the ratio of the pressure difference between Pmca and CVP and CO [RVR = (Pmca – CVP) /CO]. This formula will be used to describe venous return during transient states of imbalances (Pmca is the average pressure in the circulation and RVR is the resistance encountered to the heart).^26,27^

##### 6.1.3 Efficiency measures

Efficiency of the heart (E_h_) will be defined as the ratio of the pressure difference between Pmca and CVP and Pmca [E_h_ = (Pmca – CVP) /Pmca] (0 ≤ E_h_ ≤ 1). This equation is proposed for the measurement of heart performance, i.e., how well the heart handles the VRdP in terms of Pmca and CVP.^5,22,24^ A value ∼1 reflects a normal heart function with CVP close to 0. During the cardiac stop ejection, right atrial pressure (i.e., CVP) is equal to the Pmca, and E_h_ approaches zero.^5^

Cardiac power [Power = CO × (MAP – CVP) × 0.0022] and power output [CPO = (CO × MAP) /451] will be also calculated. Cardiac power represents the rate of energy input the systemic vasculature receives from the heart at the level of the aortic root to maintain the perfusion of the vital organs in shock states.^28^ Power efficiency (E_power_) will be defined as the ratio between the change in power and the change in Pmca [E_power_ = Δ((MAP-CVP) × CO) × 0.0022 /ΔPmca]. Whereas E_h_ is a static variable, E_power_ dynamically describes the change in cardiac power in relation to the change in power (MAP × CO) and Pmca.^29^

Volume efficiency (E_vol_) will be calculated as the ratio of the pressure difference between Pmca and right atrial pressure (i.e., CVP) and the change in Pmca [E_vol_ = Δ(Pmca-CVP) /ΔPmca] (0 ≤ E_vol_ ≤ 1). Volume efficiency conveys a dynamic variable embodying the efficiency of added fluid, vasopressor, or inotrope in terms of increase in VRdP related to increase in Pmca, and therefore CO and oxygen delivery.^29^

##### 6.1.4 Other circuit parameters

We will also calculate arterial compliance [C_art_ = SV /(SAP – DAP)], arterial resistance [R_art_ = MAP /(SV × HR)], venous compartment resistance [R_ven_ = SVR × 0.038], and effective arterial elastance (E_a_ = MAP /SV) which is an integrative measure of cardiac afterload that includes steady and pulsatile components. Dynamic arterial elastance will be calculated as the ratio of pulse pressure variation to stroke volume variation, serving as a functional measure of ventriculo-arterial coupling.

#### 6.2. Multimodal assessment of peripheral perfusion

The term “multimodal assessment” describes the systematic and comprehensive monitoring of multiple measures to assess particular patient characteristics. Because it is difficult for a single parameter to adequately reflect overall peripheral perfusion in septic patients, this multifaceted assessment has a high clinical reference value.^10^ A comprehensive multi-parameter analysis may correctly assess the association of macrohemodynamics with peripheral perfusion parameters and effectively guide treatment.

The following parameters will be evaluated every four hours for three consecutive days: (a) skin temperature; (b) lactate representing the degree of cell/tissue hypoxia; (c) ScvO_2_ representing systemic oxygen consumption; (d) oxygen extraction ratio (O_2_ER); (e) urine output, (f) venous–arterial carbon dioxide difference (Pv-aCO_2_) representing tissue perfusion and oxygenation imbalance, (g) skin mottling, (h) CRT representing microcirculatory perfusion status, (i) StO□, and (j) arterial blood glucose and interstitial fluid glucose difference (G_A−I_). These perfusion parameters and their kinetics represent many physiological mechanisms of the human body.

### 7. Outcomes

#### 7.1 Primary outcome

- Change in SOFA and SAPS II scores from baseline (enrollment) to 72 hours.

#### 7.2 Key secondary outcomes

- 28-day mortality.
- Intensive care unit and hospital length of stay.
- Number of ventilator- and vasopressor-free days at day 28.
- Lactate clearance at 6, 24, 48, and 72 hours.
- Safety outcomes, including fluid balance, incidence of pulmonary edema or respiratory failure, arrhythmias, and requirement for renal replacement therapy.

#### 7.3 Safety outcomes

- Net fluid balance at 72 hours.
- New need for renal replacement therapy within 7 days.
- Major arrhythmias requiring treatment.
- Clinician-reported adverse events related to protocol (pulmonary edema, ischemic limb events, etc.).

### 8. Data collection, monitoring, management, and schedule of assessments

Clinical data will be obtained through a review of electronic medical records and medical charts. Data analysis will be based on predefined and contemporaneously recorded measurements. The staff will be blinded to measurements until the end of the study and all data are analyzed. An independent Data and Safety Monitoring team will oversee safety, ethical, and scientific aspects of the study. The goal of the clinical data management plan is to provide high-quality data by adopting standardized procedures to minimize the number of errors and missing data, and consequently, to generate an accurate database for analysis. Remote monitoring will be performed to signal early aberrant patterns, issues with consistency, credibility, and other anomalies. Any missing and outlier data values will be individually revised and completed or corrected whenever possible. An adjudication committee, blinded to treatment allocation, will review the data to determine sepsis events and causes of death. All researchers will be trained in advance.

The general characteristics of the patients will be recorded, including demographic information, diagnosis, disease severity (e.g., SOFA and SAPS II scores —a higher score within 24 h after enrollment indicates more severe organ dysfunction^30,31^— and Acute Physiology and Chronic Health Evaluation (APACHE) II score —a higher APACHE II score indicates more severe disease, a poorer prognosis, and a higher mortality rate^32^), and tissue perfusion parameters. Patients will be subjected to standardized bedside testing at the time of enrollment (H0), H4, H8, H12, H16, H20, H24, H28, H32, H36, H40, H44, H48, H52, H56, H60, H64, H68, and H72 according to predetermined schemes.

In terms of perfusion parameters, CRT is the amount of time that passes between a physician applying the proper pressure to a patient’s fingernail bed for 15 seconds in order to precisely remove blood from the nail tip and create a crescent (whitening) underneath the nail bed and the time until the complete restoration of the nail bed’s color.^11^ To reduce measurement error, capillary refill time will be measured three times in a row by two physicians before the mean value is determined.

The skin mottling score is used to conduct a semi-quantitative evaluation of skin mottling at the patient’s bedside and represents the size of mottled regions on the patients’ knees and thighs when they lie supine with their legs stretched out and flush with their hearts.^33^ This system uses a scoring system with a range of 0 to 5; a score of 0 indicates that there is no mottling, while scores 1 through 5 indicate progressively more severe mottling. In particular, a score of 1 means that the mottling is limited to the knee’s center, whereas a score of 2 means that it extends to the kneecap’s perimeter. A score of four means that the mottling has expanded to the ends of the thigh and calf, whereas a score of three means that it is above the knee but does not extend past the mid-thigh and calf. Lastly, mottling that has extended to the ankle and groin is represented by a score of 5.

Using a central venous catheter, a central venous blood sample will be drawn from the patient, and a blood gas analyzer will measure the amount of ScvO_2_ in the blood sample. Additionally, Pv-aCO_2_ will represent the carbon dioxide difference, which is determined by subtracting the partial pressure of carbon dioxide in the arterial blood from that in the central venous blood. This information will be derived from the simultaneous collection of arterial and central venous samples.

Using a near-infrared spectroscopy (NIRS) instrument, the skeletal muscle microvascular oxygenation patterns of the right vastus lateralis will be assessed. After the area has been carefully shaved and dried, the probe will be affixed to the skin above the vastus lateralis muscle (about 12 cm above the kneecap) using adhesive tape and an elastic bandage. Oxygenated and deoxygenated hemoglobin/myoglobin concentrations (oxy-[Hb/Mb] and deoxy-[Hb/Mb], respectively) can be measured using NIRS.^34^ Additionally, tissue oxygen saturation (StO□ = oxy-[Hb/Mb]/total - [Hb/Mb]) and total heme concentration (total-[Hb/Mb] = oxy-[Hb/Mb] + deoxy-[Hb/Mb]) will be computed.^34^ The deoxy-[Hb/Mb] signal, which was measured from resting baseline to fatigue, has been regarded as a surrogate marker of fractional oxygen extraction in the microcirculation, representing the balance between oxygen delivery (DO_2_) and consumption (VO_2_).^35^

The finger-pricking technique and a standard glucose meter will be used to collect interstitial fluid glucose at the same time as arterial blood gas analysis and arterial blood glucose measurement.^36^ The interstitial fluid glucose difference will be calculated as the difference between the two measurements.

Sepsis will be diagnosed via clinical assessments and basic laboratory tests, cultures, imaging studies, and sepsis biomarkers, as indicated, in conjunction with SIRS criteria [tachycardia (heart rate >90 beats/min), tachypnea (respiratory rate >20 breaths/min), fever or hypothermia (temperature >38 or <36 °C), and leukocytosis, leukopenia, or bandemia (white blood cells >1,200/mm^3^, <4,000/mm^3^ or bandemia ≥10%)] (https://www.mdcalc.com/calc/1096/sirs-sepsis-septic-shock-criteria). ^17,18^

Sepsis-related, non-compensatory tachyarrhythmia will be managed with intravenous landiolol, titrated based on systemic hemodynamics and tissue perfusion.^6,37,38^ Macrohemodynamic evaluation will also include analysis of the systolic–dicrotic pressure difference, which may indicate a reduced rate of pressure change over time (dP/dt_max) and reflect the degree of coupling between myocardial contractility and afterload.^39^ Tissue perfusion will be assessed using the previously described multi-parameter analysis of peripheral perfusion.

### 9. Safety monitoring

Safety monitoring will be overseen by a Data Safety and Monitoring Board (DSMB), which will review safety data at 25%, 50%, and 75% of enrollment, as well as on an ad hoc basis as needed. The DSMB will have the authority to recommend pausing or stopping the trial for safety concerns, such as a significant increase in 28-day mortality or major adverse events in the intervention arm, or for futility based on predefined boundaries.

### 10. Sample size and predefined statistical analysis plan

The trial is powered to detect a 2.0-point between-group difference in change of SOFA and SAPS II from baseline to 72 hours, assuming a common SD = 3.0, two-sided α = 0.05, and 90% power. Using standard sample-size formulas, this requires ≈47.3 subjects per group; rounding yields 48 per group. Allowing for 10% attrition, the target enrollment is 54 per group (total N = 108). Sensitivity scenarios for smaller effect sizes and larger variance are presented in Appendix A.

Change in SOFA and SAPS II will be analyzed using linear mixed-effects models with treatment × time interaction, adjusting for baseline score and stratification variables (site, APACHE II category). A hierarchical testing strategy (SOFA first; if significant at α = 0.05, then SAPS II) will be used to preserve familywise error. Interim analyses at 25%, 50%, and 75% enrollment will be conducted under DSMB oversight using an O’Brien–Fleming spending function. Missing data will be handled primarily via LMM (MAR assumption) with multiple imputation and MNAR sensitivity analyses as prespecified.

All statistics will be performed using R software version 4.0.5.

### 11. Trial governance and timeline

Trial governance and timeline are structured to ensure rigorous scientific oversight and efficient study conduct. The Trial Steering Committee is responsible for supervising the scientific integrity of the trial. The study will be conducted at a coordinating site, Tzaneio General Hospital, along with 2–4 additional ICUs to facilitate timely patient enrollment. The anticipated enrollment period is 10–12 months, although this may vary depending on recruitment rates.

### 12. Ethics and dissemination

The study protocol will be approved by the Ethics Committee of Tzaneio General Hospital and the study will be conducted in accordance with the Declaration of Helsinki, the International Conference on Harmonization Good Clinical Practice (ICH-GCP) guidelines, and applicable local regulatory requirements. Written informed consent will be obtained from each patient or, when necessary, from a legally authorized representative prior to the initiation of any study procedures. The final study dataset will be made available to the investigators and retained for five years following trial completion. Study findings will be disseminated through presentations at national and international scientific conferences and publication in peer-reviewed journals, with anticipated reporting between 2026 and 2027.

## III. DISCUSSION

Effective sepsis management requires early detection and timely intervention, as delays substantially increase morbidity and mortality among critically ill patients.^40^ Conventional systemic hemodynamic parameters may not reliably reflect tissue-level hypoperfusion, leading to the phenomenon of hemodynamic incoherence, in which macrocirculatory variables appear adequate while microcirculatory flow and cellular metabolism remain compromised. To address this challenge, the PRISM trial integrates advanced hemodynamic indices with a structured, multimodal assessment of tissue perfusion, aiming to improve both diagnostic accuracy and therapeutic guidance.

Clinical examination remains essential in sepsis. Peripheral skin temperature gradients, particularly between the core and extremities, reflect sympathetic vasoconstriction; a central-to-toe temperature difference >4°C has been associated with impaired perfusion and adverse outcomes.^41^ Capillary refill time >3 seconds correlates with elevated lactate and higher 28-day mortality.^42^ Likewise, mottling assessed with a semi-quantitative score predicts prognosis; a mottling score ≥3 at 6 hours was linked to >60% mortality compared to <10% for a score of 0.^33^ Urine output, though nonspecific, reflects renal perfusion, with sustained oliguria independently associated with ICU mortality.^43^

In addition, the importance of advanced hemodynamic assessment in sepsis management is increasingly recognized. Traditional parameters, such as MAP and CO, capture only limited aspects of circulatory function and may appear normal despite significant underlying circulatory abnormalities. Incorporating measures that reflect determinants of venous return and cardiovascular efficiency allows for a more integrated, systems-level evaluation of cardiovascular performance.^5,22,24^ These indices provide mechanistic insights into the interplay between preload, afterload, and cardiac pump function, revealing states of circulatory failure that conventional metrics may fail to detect.

Of note, sepsis-associated, non-compensatory tachyarrhythmia —driven by sympathetic overstimulation and autonomic dysfunction— can lead to myocardial injury and worsen patient outcomes. Although β-blockers have demonstrated efficacy in this context, clinical debate persists regarding the optimal timing and intensity of heart rate control, with concerns about potential adverse effects such as hypotension. In the PRISM trial, sepsis-related, non-compensatory tachyarrhythmia will be treated with an intravenous ultra–short-acting selective β-blocker (landiolol), titrated according to systemic hemodynamics and tissue perfusion.^6,37,38^ Integrating these physiologically informed metrics into bedside care can improve resuscitation precision and align therapeutic targets with the patient’s true circulatory reserve and oxygen delivery capacity, thereby optimizing heart rate management.

Biochemical and oxygen-derived markers complement physical examination and hemodynamic assessment. Serum lactate remains a cornerstone biomarker, with elevated levels (>2 mmol L^-1^) predicting worse outcomes and failure of clearance (>10% within 6 hours) indicating poor prognosis.^44^ Yet lactate may rise from adrenergic stimulation or impaired clearance, underscoring the need for additional indices. Central venous oxygen saturation values <70% may reflect inadequate delivery, whereas high ScvO_2_ with rising lactate suggests impaired extraction.^45^ The O_2_ER provides further granularity: values >30% in the context of hypotension or organ dysfunction imply inadequate perfusion.^46^ The Pv–aCO_2_ is a sensitive marker of low-flow states; values >6 mmHg are associated with reduced CO and poor outcomes, even with normal ScvO_2_, while persistent elevation after resuscitation nearly doubles 28-day mortality.^47,48^ The Pv–aCO_2_/C(a–v)O_2_ ratio >1.4 has been proposed as a marker of anaerobic metabolism.^49^

Near-infrared spectroscopy will allow continuous assessment of StO_2_. Septic patients typically exhibit lower baseline StO_2_, with levels <75% linked to higher mortality.^50^ Impaired StO_2_ recovery after vascular occlusion testing independently predicts adverse outcomes,^51^ while meta-analysis confirms its prognostic utility.^52^ Similarly, impaired glucose utilization at the tissue level may be detected by widening arterial–interstitial glucose gradients, reflecting microvascular dysfunction and cellular metabolic stress.^36,53-55^

The PRISM trial carries significant implications. First, bedside clinicians will gain actionable insights by integrating structured perfusion assessments with advanced systemic hemodynamics. Second, individualized resuscitation strategies could prevent occult hypoperfusion from progressing to organ failure. By linking predefined abnormal thresholds (e.g., CRT >3 sec, StO_2_ <75%, Pv–aCO_2_ >6 mmHg, rising lactate, mottling score ≥2) to a structured treatment algorithm, the trial ensures resuscitation is both evidence-based and physiologically coherent, while preserving therapeutic discretion for the treating clinician to enhance real-world applicability. Notably, the trial supports the targeted titration of non-compensatory tachyarrhythmia with an ultra–short-acting selective β-blocker, guided by key physiological parameters to optimize hemodynamic stability without compromising perfusion. Finally, by using SOFA and SAPS II as co-primary outcomes, PRISM assesses not only organ-specific dysfunction but also overall disease severity and prognosis.

Potential limitations include the open-label design, variability in protocol adherence, and a sample size primarily powered for changes in SOFA and SAPS II rather than mortality. Nevertheless, blinded outcome adjudication, rigorous measurement protocols, and a predefined statistical analysis plan strengthen the robustness and reproducibility of the findings.

In conclusion, the PRISM trial pioneers a physiology-driven, multimodal approach to septic shock, aligning systemic hemodynamics with tissue perfusion to bridge the gap between macrocirculatory stabilization and microcirculatory recovery, ultimately aiming to reduce organ dysfunction and improve survival.

## Conflicts of interests

The author has no conflicts of interest.

## Data availability

Data will be made available upon request after publication through a collaborative process. Researchers should provide a methodically sound proposal with specific objectives in an approval proposal.

## Acknowledgements

Nothing to acknowledge.

## Author contributions

AC contributed to study design, protocol draft, review and editing of the manuscript.

## Competing interests

None declared.

## Funding

None.

## Appendix A.

## Sample size and predefined statistical analysis plan (90% power)

### Sample size — primary endpoints

Primary endpoints: change in SOFA score and change in SAPS II score from baseline to 72 hours.

Primary hypothesis: personalized management guided by multimodal circulatory and perfusion monitoring produces a greater reduction in organ-dysfunction scores (SOFA and/or SAPS II) at 72 h compared with standard care.

Assumptions used for sample size calculations:

- Target between-group difference (effect size), δ = 2.0 points (difference in change from baseline to 72 h).
- Common standard deviation of change scores, σ = 3.0.
- Two-sided type I error (familywise) α = 0.05.
- Power = 90% (1 − β = 0.90).
- Anticipated loss to follow-up /missing primary endpoint data: 10%.
- Randomization stratified by site and baseline APACHE II category (≤15 vs >15).

Numeric calculation (two-sample approximation for transparency) n per group = 2 × (Z1−α/2 + Z1−β)^2^ × σ^2^ /δ^2^

Z1−α/2 = 1.96 (two-sided α = 0.05)

Z1−β = 1.2816 (power = 90%)

With δ = 2 and σ = 3: n per group ≈ 47.3

Round up:

- 48 subjects per group (pre-attrition).
- Allowing for 10% attrition: 54 subjects per group.
- Total target enrollment = 108 patients (54 per arm).

### Sensitivity scenarios (90% power, α = 0.05)

- δ = 1.5, σ = 3 → 95 per group, total 190.
- δ = 2.5, σ = 3 → 35 per group, total 70.
- δ = 2.0, σ = 4 → 95 per group, total 190.

The table below shows required sample sizes for combinations of effect sizes (δ) and standard deviations (σ) under the assumptions: two-sided α=0.05, power=90%, and 10% attrition. Calculation formula: n per group = 2 × (Z_{1−α/2} + Z_{1−β})^2 × σ^2 /δ^2, with Z_{1−α/2}=1.96 and Z_{1−β}=1.2816.

| Effect size ($\delta$) | SD ($\sigma$) | n per group<br>(calc) | n per group<br>(round up) | n per group<br>(with 10%<br>attrition) | Total N (with<br>attrition) |
| --- | --- | --- | --- | --- | --- |
| 1.5 | 3.0 | 84.06 | 85.0 | 95.0 | 190.0 |
| 2.0 | 3.0 | 47.29 | 48.0 | 54.0 | 108.0 |
| 2.5 | 3.0 | 30.26 | 31.0 | 35.0 | 70.0 |
| 1.5 | 4.0 | 149.45 | 150.0 | 167.0 | 334.0 |
| 2.0 | 4.0 | 84.06 | 85.0 | 95.0 | 190.0 |
| 2.5 | 4.0 | 53.8 | 54.0 | 60.0 | 120.0 |

### Multiplicity and primary endpoint strategy

Two co-primary measures (SOFA and SAPS II). A hierarchical gatekeeping strategy will be used to preserve familywise error without inflating sample size:

1. Test change in SOFA at two-sided α = 0.05.
2. Only if SOFA is significant, test change in SAPS II at α = 0.05.

### Predefined statistical analysis plan

‐ Intention-to-treat (ITT) as primary analysis; per-protocol (PP) and as-treated as sensitivity.
‐ Primary analysis: linear mixed-effects models (LMM) with treatment × time interaction, adjusting for baseline and stratification variables. Supportive ANCOVA for 72 h values.
‐ Secondary outcomes: logistic regression and Cox models for 28-day mortality; competing-risk or negative binomial models for LOS; LMM for lactate clearance; logistic regression for safety events.
‐ Interim analyses at 25%, 50%, and 75% enrollment with O’Brien–Fleming alpha-spending function under DSMB oversight.
‐ Missing data: handled by LMM (MAR assumption) and multiple imputation (MICE with 20 datasets) as sensitivity. MNAR sensitivity via tipping-point analyses.
‐ Subgroup analyses: exploratory, by APACHE II category, lactate level, sepsis source, age, and baseline cardiac function. Interaction tests reported with caution.

All statistics will be performed using R software version 4.0.5.

## Notes

### Competing Interest Statement

The authors have declared no competing interest.

### Clinical Trial

The trial will be prospectively registered at ClinicalTrials.gov

### Funding Statement

This study did not receive any funding

### Author Declarations

The study protocol will be approved by the Ethics Committee of Tzaneio General Hospital, Piraeus, Greece.

